# INFLAMMATORY MARKERS AND BODY MASS INDEX AMOUNG HISPANIC CHILDREN

**DOI:** 10.1101/2023.07.21.23293009

**Authors:** Henry Lang, Elaine N. Loudermilk, W. Andrew Clark, Jo-Ann Marrs, T. Andrew Joyner, Liang Wang, Kathryn S. Gerber, Arsham Alamian

## Abstract

**Background and Objectives:** Body mass index (BMI) is inversely proportional with adiponectin levels among adults, while insulin, C-reactive protein (CRP), interleukin 6 (IL-6), resistin, and tumor necrosis factor-alpha (TNF-α) have been linked with elevated BMI. The role and relation of these biomarkers with BMI among Hispanic pediatric populations are less known. Thus, the objective of this cross-sectional study was to examine the association of inflammatory markers with the odds of overweight/obesity while controlling for several sociodemographic factors among a Hispanic youth population of Northeast Tennessee.

**Methods:** Height, weight, demographic information, and blood samples were collected from 107 Hispanic children aged 2 to 10 years recruited at a large community health center in 2015-2016 in Northeast Tennessee. Data for this research were accessed and analyzed in 2022. Multivariable logistic regression was conducted to assess the relations between adiponectin, insulin, resistin, CRP, TNF-α, and IL-6, and overweight/obesity vs. having a healthy (normal) weight.

**Results:** Adiponectin levels were significantly lower among overweight/obese Hispanic children (p = 0.0048) compared to healthy weight children. The odds of overweight/obesity decreased by 4% for every one-unit increase in serum adiponectin. Insulin levels were significantly higher among overweight/obese Hispanic children (p = 0.0048) compared to healthy weight children (*p=*0.0008). The odds of overweight/obesity increased by 7% for every one-unit increase in serum insulin. Resistin, IL-6, TNF-α, and CRP were not significantly associated with overweight/obesity in this population.

**Conclusion:** Adiponectin behaves similarly in Hispanic youth as it does in other pediatric populations, possibly making it a valuable marker when examining metabolic health status in this population.

## Introduction

The prevalence of obesity among youth ages 2 to 19 years old in the United States (U.S.) increased from 17.7% in 2011-2012 to 21.5% in 2017-2020. [1] Hispanic populations and groups identified as having a low socioeconomic status (SES) are at higher risk for obesity.[2–4] Increasing prevalence of obesity in the children is concerning due to the shared relationship of childhood and adult obesity; children with obesity are at a greater risk to have obesity during adulthood than children with normal weight.[5,6] Obesity in turn increases risk for metabolic conditions, atherosclerosis, stroke, sleep apnea, respiratory disorders, and certain cancers as an individual ages.[5,6] The development of atherosclerotic lesions on vascular walls also leads to atherosclerosis which is a major contributor to cardiovascular disease.[7] The atherogenic process can begin in early childhood and remain throughout life.[7] Risk for developing atherosclerosis and additional complications due to obesity can be monitored by tracking specific physiological biomarkers.

Previous studies have shown body mass index (BMI) to be inversely proportional with adiponectin levels among adults and children.[8,9] Increased levels of adiponectin have been associated with a reduction in glycogenolysis, due to increased uptake of glucose by adipose tissue,[10] and hepatic gluconeogenesis. The downregulation of hepatic gluconeogenesis is coupled with an increase in fatty acid β-oxidation, suggesting an increase in energy expenditure.[11] Insulin is a negative regulator of gluconeogenesis and chronically elevated levels are associated with a reduction in adiponectin.[12] Treatment with adiponectin in rats demonstrated an increase in insulin-stimulated glucose uptake in adipose tissue through the upregulation of adenosine monophosphate protein kinase (AMPK), reducing the phosphorylation of p70 S6 kinase, which acts as an inhibitor.[13] Conversely, C-reactive protein (CRP), interleukin 6 (IL-6), resistin, and tumor necrosis factor-alpha (TNF-α) are associated with elevated BMI and inflammation. These markers have also been linked to increased risk for developing atherosclerotic lesions and have been found to be inversely proportional to adiponectin.[14–16] The relationship between adiponectin and BMI among children has been explored, highlighting a negative correlation between adiposity and adiponectin.[17,18] However, there remains a gap in the literature regarding the relationship of adiponectin, CRP, IL-6, resistin, and TNF-α with BMI among Hispanic children. Thus, the objective of this study was to examine the association between inflammatory markers and the likelihood of being overweight/obese while controlling for sociodemographic factors within Hispanic youth in Northeast Tennessee.

## Methods

### Participants

Secondary analysis of height, weight, demographic information, and blood samples from 114 Hispanic children aged two to ten years recruited in 2015-2016 at a large community health center in Northeast Tennessee, as part of a larger study on metabolic syndrome,[19] were completed. Data for this article were accessed for research purposes and analyzed from September to December 2022. Of the 114 children considered, 1 was excluded due to having a BMI below the 5^th^ percentile (i.e., underweight), 2 had missing data on maternal education, and 4 had missing information on TNF-α, resistin and adiponectin. The final analytic sample size comprised of 107 children. Methods for the collection of data used are mentioned elsewhere in a study conducted by Alamian et al.[19] The authors did not have access to information that could identify individual participants during or after data collection.

This study received approval from the East Tennessee State University Human Subject Research Ethics Committee (IRB#: 0414.16s). Written consent was obtained from all study participants.

### Dependent variable

The outcome of interest, BMI status (obese defined as overweight/obese vs. healthy weight, according to 2000 CDC growth charts),[20] was treated as a categorical variable.

### Independent variables

The variables of interest included adiponectin (µg/mL), insulin (uIU.L), resistin (pg/mL), TNF-α(pg/mL), CRP (mg/dL), and IL-6 (pg/mL). Analysis for adiponectin (171A7002M), insulin and resistin (171A7001M), and TNF-α and IL-6 (171A7002M) were performed using Bio-Rad Bio-Plex Mag-Pix according to provider procedures. East Tennessee State University Clinical Laboratory: an accredited reference lab (Center for Medicare & Medicaid Services Clinical Laboratory, certification number 44D0659180) was utilized for analysis of serum CRP.

### Covariates

Covariates included child age, child sex (male or female), maternal education level (high school graduation or more vs. less than high school education), maternal marital status (married versus other). Child age was reported as a continuous variable in years.

### Statistical analysis

Descriptive statistics (frequencies and percentages for categorical variables, and means and standard deviations for continuous variables) were performed to describe the data as appropriate. Pearson chi-squared test and independent *t*-tests were conducted to examine differences in percentages (for categorical variables) and means (for continuous variables) by weight status (overweight/obese versus healthy weight status). Simple and multiple logistic regression analyses were then performed to assess the strength of association between variables of interest and being overweight/obese versus healthy weight among participants in this population. Multiple logistic regression models controlled for the potential effects of covariates (age, sex, maternal education, maternal marital status). Statistical analysis was completed via statistical analyst system (SAS, version 9.4; SAS Institute, Cary, NC).

## Results

The majority of the study sample were female (54.21%) with an average age of 6.62 years (SD: 2.74 years) as seen in Tables 1 and 2. Almost half of the sample were classified as overweight/obese (44.86%) with the remaining sample having a healthy weight status (55.14%). Slightly over half of mothers had less than a high school education (56.07%) and most were married (78.50%). A greater percentage of males (48.98%) were overweight/obese compared to females (41.38%), although this difference was not statistically significant. There were no statistically significant differences in weight status by maternal education or marital status (P > 0.05). Average levels of biomarkers were as follows: Adiponectin, 24.27 µg/mL (SD=13.85); insulin, 16.03 uIU.L (SD=17.46); resistin, 5510.23 pg/mL (SD=3817.13); TNF-α, 8.40 pg/mL (SD=17.00); CRP, 2.35 mg/dL (SD: 4.86); and IL-6, 3.23 pg/mL (SD=8.68).

**Table 1.** Sociodemographic characteristics of the Hispanic pediatric sample by healthy and overweight/obese weight status (n = 107).^a^.

|  | **N (%)**<br>**Total** | **Healthy**<br>**Weight** | **Overweight/Obese** | **P-**<br>**value<sup>b</sup>** |
| --- | --- | --- | --- | --- |
| **n, %, Total** | 107<br>(100.00) | 59 (55.14) | 48 (44.86) |  |
| **Sex, n (%)** |  |  |  | 0.4310 |
| Male | 49 (45.79) | 25 (51.02) | 24 (48.98) |  |
| Female | 58 (54.21) | 34 (58.62) | 24 (41.38) |  |
| **Maternal education, n (%)** |  |  |  | 0.6711 |
| Less than high school education | 60 (56.07) | 32 (53.33) | 28 (46.67) |  |
| High school graduate or more | 47 (43.93) | 27 (57.45) | 20 (42.55) |  |
| **Maternal marital status, n (%)** |  |  |  | 0.2044 |
| Married | 84 (78.50) | 49 (58.33) | 35 (41.67) |  |
| Other | 23 (21.50) | 10 (43.48) | 13 (56.52) |  |

a. Data from a study of metabolic syndrome in Hispanic children in Johnson City, TN [19].
b. P-value from the chi-squared test.

**Table 2.** Mean (SD) for child age and selected biomarkers by healthy weight and overweight/obese weight status among the Hispanic pediatric sample (N=107).^a^.

|  | Healthy Weight (N=59) |  | Overweight/Obese (N=48) |  |  |
| --- | --- | --- | --- | --- | --- |
|  | Mean | Std. Dev. | Mean | Std. Dev. | P-value<sup>b</sup> |
| Child age (years) | 6.39 | 2.75 | 6.90 | 2.73 | 0.3446 |
| Adiponectin (ug/mL) | 27.53 | 15.37 | 20.27 | 10.56 | 0.0048 |
| Insulin (uIU.L) | 10.61 | 9.94 | 22.70 | 22.00 | 0.0008 |
| Resistin (pg/mL) | 4916.50 | 2387.40 | 6240.00 | 4984.10 | 0.0961 |
| TNF-alpha (pg/mL) | 8.18 | 13.23 | 8.67 | 20.88 | 0.8877 |
| C-reactive protein (mg/dL) | 2.33 | 5.48 | 2.38 | 4.01 | 0.9626 |
| Interleukin 6 (pg/mL) | 3.73 | 10.47 | 2.62 | 5.84 | 0.4888 |

a. Data from a study of metabolic syndrome in Hispanic children in Johnson City, TN [19].
b. P-value from t-test.

Results from the independent *t*-tests for differences in biomarkers by weight status are shown in Table 2. Adiponectin levels were significantly lower among overweight/obese children compared to healthy children (*p* = 0.0048). Insulin levels were significantly higher among overweight/obese children compared to healthy children (*p=*0.0008). No other significant differences were observed between overweight/obese and healthy children when considering other biomarkers or the child’s age.

Simple logistic regression (SLR) and multiple logistic regression (MLR) results are presented in Table 3. In SLR models, both adiponectin and insulin were significantly associated with being overweight/obese. For every one-unit increase in serum adiponectin, the child’s odds of overweight/obesity decreased by 4% [Odds Ratio (OR): 0.96, 95% confidence interval (95% CI): 0.93-0.99, P=0.0084). In contrast, every one-unit increase in insulin resulted in 7% increased chance of overweight/obesity [OR: 1.07, 95% CI: 1.03-1.11, 0.0017. MLR results corroborated the SLR findings: both adiponectin and insulin were found to be significantly associated with overweight/obesity. For every one-unit increase in serum adiponectin, the odds of overweight/obesity decreased by 4% (OR: 0.96, 95% CI: 0.93-0.99) while holding all other variables constant. For every one-unit increase in insulin, the odds of overweight/obesity increased by 7% (OR): 1.07, (95% CI): 1.02-1.13. Child age, child sex, maternal education and marital status, and other biomarkers (resistin, IL-6, TNF-α, and CRP) were not significantly associated with overweight/obesity.

**Table 3.** Simple and multiple logistic regression examining the relationship between biomarkers and overweight/obesity in a Hispanic pediatric sample from Northeast TN (N=107)

| Variables | SLR |  |  | MLR |  |  |
| --- | --- | --- | --- | --- | --- | --- |
|  | OR | 95 % CI | P-value | OR | 95 % CI | P-value |
| Adiponectin (ug/mL) | 0.96 | 0.93-0.99 | 0.0084 | 0.96 | 0.93-0.99 | 0.0104 |
| Insulin (uIU/L) | 1.07 | 1.03-1.11 | 0.0017 | 1.07 | 1.02-1.13 | 0.0052 |
| Resistin (ug/mL) | 1.00 | 1.00-1.00 | 0.1000 | 1.00 | 1.00-1.00 | 0.0477 |
| TNF-alpha (pg/mL) | 1.00 | 0.98-1.02 | 0.8813 | 1.01 | 0.95-1.08 | 0.6748 |
| C-reactive protein (mg/dL) | 1.00 | 0.93-1.08 | 0.9632 | 0.99 | 0.90-1.09 | 0.8879 |
| Interleukin 6 (pg/mL) | 0.98 | 0.93-1.04 | 0.5235 | 0.91 | 0.75-1.09 | 0.2867 |
| **Child sex** |  |  |  |  |  |  |
| Female vs male | 0.735 | 0.34-1.59 | 0.4314 | 0.71 | 0.28-1.82 | 0.3041 |
| **Child age** (years, continuous) | 1.28 | 0.58-2.83 | 0.3415 | 0.82 | 0.31-2.13 | 0.8268 |
| **Maternal education level** |  |  |  |  |  |  |
| High school education or less | Reference |  |  | Reference |  |  |
| High school graduate or more | 0.85 | 0.39-1.83 | 0.6712 | 0.67 | 0.27-1.70 | 0.4310 |
| **Maternal marital status** |  |  |  |  |  |  |
| Other vs married | 1.82 | 0.72-4.62 | 0.2077 | 1.58 | 0.53-4.70 | 0.4592 |

## Discussion

Results of this study indicate that both adiponectin and insulin levels are potentially significant predictors of childhood overweight/obesity. Adiponectin is still protective against increased weight even when accounting for insulin. Results from this study confirm findings from similar studies that demonstrated significantly lower adiponectin levels in Italian and Pima Native American children with overweight/obese classification using BMI.[17,18] The repeated findings of low adiponectin in overweight/obese children may be related to the relationship between adiponectin and insulin. Depressed adiponectin concentration and chronically elevated insulin have been demonstrated in the pathogenesis of insulin resistance, and therefore metabolic syndrome.[21,22] The depressed levels of adiponectin found in overweight/obese children, coupled with participant age and the lack of significant differences between groups in the other biomarkers examined, suggests adiponectin levels may be an early indicator to predict risk of developing insulin resistance and metabolic syndrome in children. While BMI has also been used as an early predictor of metabolic syndrome in children,[23] research output is increasing regarding metabolically healthy (MHO) vs metabolically unhealthy (MUO) individuals with obesity, shifting the focus of intervention trials towards improved metabolic health status as the desired outcome rather than weight loss. Alterations in BMI are not required following life-style intervention for metabolic health to improve.[24,25] Adiponectin may be a valuable variable to monitor when tracking the effects an individual’s BMI may have on their metabolic health (i.e., if an individual has high adiponectin and elevated BMI, there may be less of concern for weight-loss intervention).[26] Longitudinal studies controlling for BMI while monitoring insulin sensitivity and adiponectin in children are needed to confirm this hypothesis.

The lack of significance of resistin, CRP, IL-6, and TNF-α between the normal BMI and overweight/obese groups may be due, in part, to the pathological progression of obesity. The infiltration of inflammatory cells into adipocytes is thought to occur after a decrease in adiponectin,[27] suggesting that duration of overweight/obesity is a key factor in the progression of chronic inflammation. The participants in the current study may not have had overweight/obesity for a sufficient length of time to illicit the inflammatory consequences typically associated with obesity in adulthood.[28] Previous research regarding resistin levels in the adult population is mixed with some researchers finding greater values in individual’s with obesity compared to those with healthy weight, whereas other findings demonstrated significantly greater resistin levels in populations with obesity.[29,30] Other literature has demonstrated the capabilities of CRP, IL-6, and TNF-α to predict overweight/obesity.[31] The findings from the current study indicates adiponectin may be an earlier predictor for the risk of developing insulin resistance compared to CRP, IL-6, and TNF-α in a pediatric population. Further research in the Hispanic pediatric population is required to confirm the findings of the current study.

The adjusted analysis in this study suggests a negative association between maternal education and BMI, however findings were not significant. Previous research has demonstrated parental education to be a predictor of child overweight/obese status in countries with a high economic status. Lê-Scherban and colleagues (2021) revealed childhood obesity prevalence to be greater among children whose parents did not obtain a high school education compared to children with parents who had a college degree.[32] Ogden and colleagues (2018) found childhood obesity prevalence to be significantly lower in Hispanic US households when the head of the household was a college graduate compared to a high school graduate or less. [33]The current study examined the effect of childhood obesity between non-high school graduates to high school graduates or higher, which may elicit a smaller effect on childhood obesity when compared to the effect of non-high school graduates to college graduates.

This study is limited by a small sample size, possibly leading to a lack of statistical power. However, significance was still found despite the limited sample size indicating the relationships uncovered should be studied further. Furthermore, the cross-sectional study aided in determining associations to allow for future studies to expand on the present findings. The current study alone, however, is not enough to determine causation, thus longitudinal research are warranted. Finally, the sample population is of a specific region in Tennessee where obesity is known to be prevalent,[34] thus additional studies should seek to examine similar populations in urban, suburban, and rural settings to improve external validity.

## Conclusion

In summary, this study revealed adiponectin behaves similarly in Hispanic children as it does among adults potentially making it a valuable marker when examining health status among Hispanic pediatric populations. CRP, IL-6, and TNF-α may not be relevant markers to predict overweight/obese status in Hispanic youth, as these markers may not become consistently elevated until an individual has maintained overweight/obese status for an extended length of time. Longitudinal studies are needed to confirm the findings relating Hispanic youth BMI to adiponectin, CRP, IL-6, and TNF-α. Additional research using larger sample sizes is needed to confirm the relationship between biomarkers and BMI among Hispanic youth.

## Data Availability

N/A

## Notes

### Competing Interest Statement

The authors have declared no competing interest.

### Funding Statement

AA and WAK received a diversity research grant (E210029) from the Tennessee Board of Regents (https://www.tbr.edu/) in support of this work. The funder had no role in study design, data collection and analysis, decision to publish, or preparation of the manuscript. The opinions or assertions herein are the views of the authors and do not necessarily reflect the official policy or position of the U.S. Air Force, the Department of Defense, or the U.S. Government.

